# Comorbidity Patterns and Associated Factors of thyroid related disorders: A community-based cross-sectional study in Chinese population

**DOI:** 10.1101/2025.04.28.25326543

**Authors:** Yanru Zhao, Haobiao Liu, Ziyuan Gao, Xin Zhao, Weiwei Wang, Hui Guo, Ying Yang, Congying Yang, Xingjun Liu, Meng Li, Xi Ding, Yushi Sun, Tianxiao Zhang, Bingyin Shi

**Author notes:** Authors contributed equally to this manuscript (Yanru Zhao, Haobiao Liu). ***Correspondence*:** Tianxiao Zhang, Tang Scholar Department of Epidemiology and Biostatistics, School of Public Health, Xi’an Jiaotong University Health Science Center, Xi’an, Shaanxi, People’s Republic of China. Mailing Address: #76 Yanta West Rd, Xi’an, Shaanxi Province, People’s Republic of China, Postal Code: 710061., Bingyin Shi Department of Endocrinology, The First Affiliated Hospital of Xi’an Jiaotong University, Xi’an 710061, People’s Republic of China.; 86-13-700298366 Mailing Address: #277 West Yanta Road, Xi’an, Shaanxi, People’s Republic of China, Postal Code: 710061. ***Email address for each co-author:*** Yanru Zhao, Haobiao Liu, Ziyuan Gao, Xin Zhao, Weiwei Wang, Hui Guo, Ying Yang, Congying Yang, Xingjun Liu, Meng Li, Xi Ding, Yushi Sun, Tianxiao Zhang, Bingyin Shi.

## Abstract

This study aims to explore the patterns and associated factors of thyroid related disease comorbidity to provide a novel insight for comorbidity prevention and management. After multi-stage stratified cluster random sampling, a total of 2272 participants were recruited, and relevant data were collected through door-to-door visits and the standardized questionnaire, anthropometric measurements and biochemical examinations. Logistic regression analysis was utilized to explore potential influences, and least absolute shrinkage and selection operator (LASSO) regression was applied to control for multicollinearity between variables. We found that there were 888 individuals with thyroid disease, yielding prevalence rates ranging from 0.35% to 17.96% for thyroid related disorders. Of these, 555 enrollees had concomitant diseases and the number of comorbidity was dominated by two conditions, with subclinical hypothyroidism and hyperlipidemia being the top ranking. Female sex acted as a protective factor for thyroid disease comorbidity, while advancing age, familial history of diabetes, large waist circumference, and fast heart rate were all substantial risk factors. The risk of comorbidity rose steeply with growing waist circumference. Appropriate measures should be taken to reduce the occurrence of comorbidity and the burden of disease.

## Introduction

The thyroid gland, a butterfly-shaped endocrine organ nestled in the neck, is charged with critical tasks in the human body, chiefly regulating energy balance and metabolism ^1^. The thyroid hormone it secretes has far-reaching effects on growth, development, and metabolic functions. However, when this vital gland malfunctions, a host of disorders may manifest, bearing significant repercussions on the global healthcare system. The popular cases of thyroid conditions include hyperthyroidism, hypothyroidism, thyroid nodule, and thyroiditis ^2^, which may be spurred on by numerous factors, including nutritionally deficient iodine status ^3,4^, genetic susceptibility ^5^, advancing age ^2^, sex ^6^, ethnicity ^7^, cigarette smoking ^8^, alcohol consumption ^9^, or exposure to environmental pollutants ^10^. In recent years, resolute awareness of healthcare, along with improved living conditions and advanced diagnostic techniques, have caused an upward trend in the prevalence of thyroid conditions ^11^. As a vastly common endocrine disease conglomerate, thyroid problems are now becoming a looming challenge to global wellness ^11^. Besides, the concomitant pathologies of thyroid disorders pose escalating concern as their incidence keeps increasing.

Thyroid diseases are multifaceted and often coincide with a myriad of health problems, such as dyslipidemia and diabetes. Numerous studies have provided evidence for an intricate connection between thyroid dysfunction and lipid profiles, revealing that low TSH levels can result in high total cholesterol (TC) and LDL-C levels in hypothyroid patients, whereas hyperthyroidism results in the opposite effect ^12^. These findings suggest that TSH plays a crucial role in the modulation of lipid metabolism. A study tracking a group of healthy adults, without diabetes mellitus, for 6 years found that an increase of serum TSH levels by 1 mU/L could potentially increase their risk of acquiring the disease by several fold ^13^. This discovery underscores the integral correlation between TSH levels, the progression of diabetes mellitus, and the emergence of insulin resistance. Furthermore, extensive epidemiological research has established a link between thyroid disorders and a range of chronic diseases, including hypertension, hyperlipidemia, and diabetes, suggesting the possible existence of shared etiologies among these conditions ^14,15^. Due to the prevalence of environmental and social factors as triggers for chronic diseases, they tend to exhibit multiple comorbidities. For instance, unhealthy lifestyle choices like sedentary behavior and smoking can cause these diseases and lead to the development of thyroid disorders underpinned by comorbidities. Despite the evidence supporting comorbidities of thyroid-related diseases, research has mainly focused on the association of single conditions with these disorders, warranting further investigation of their intricate connections. Based on this, exploring the comorbidity patterns of thyroid-related diseases is of critical importance.

Hence, our study aims to unravel the complex web of comorbidities related to thyroid diseases. By examining the associated factors, we aim to provide critical insights that could prove invaluable in the prevention and management of comorbidities. Through this research, we strive to not only expand our understanding of thyroid-related diseases but also pave the way for novel approaches in treating and preventing comorbidities.

## Materials and Methods

### Study Participants

This study is a sub-project of the Thyroid disorders, Iodine status and Diabetes Epidemiological survey (TIDE study) in Xi’an, Shaanxi Province, led by China Medical University, details of which can be found elsewhere ^16^. To ensure representative sampling, we implemented a multi-stage stratified cluster random sampling approach and selected two districts, Yanta District and Chang’an District, in the region (**Supplementary Fig. S1**). To further ensure the integrity of our results, we randomly chose two communities from each district, and study participants were recruited based on strict inclusion criteria. We only considered individuals who were 18 years of age or older, of Han ethnicity, had lived in the community for a minimum of 5 years, and were not pregnant women. A total of 2469 individuals were initially recruited, but we applied stringent exclusion criteria that included recent iodine-containing contrast exams or medication and a requirement for complete information, which resulted in 2272 study participants met all the eligibility requirements (**Supplementary Fig. S2**). All participants provided their written informed consent, and our procedures were approved by the Medical Ethics Committee of The First Affiliated Hospital of China Medical University. All methods were carried out in accordance with the Declaration of Helsinki.

### Data collection and measurement

To obtain accurate and standardized data, our team of highly trained technicians carried out a comprehensive set of measurements and examinations on study participants. Utilizing a standard questionnaire, we gathered data on demographic and social characteristics (such as residential area, sex, age, ethnicity, education, occupation, and annual income), iodine nutritional status (such as salt consumption, intake of iodine-containing drugs), lifestyle (such as smoking), and medical history.

During the physical examinations, participants were asked to wear loose clothing and remove their shoes and hats for accurate height and weight measurements. Waist circumference was also measured using a tape measure at the level of the navel. Blood pressure was monitored on the participants’ right upper limb using a high-tech electronic sphygmomanometer (Omron HEM-7430) after a 10-minute period of quiet rest, with each measurement performed twice, five minutes apart. We performed thyroid ultrasounds using a uniform portable instrument (LOGIQ 100 PRO, GE, Milwaukee, USA, with 7.5 MHz linear transducers) to assess the participants’ thyroid function.

To evaluate blood markers, we collected fasting blood samples the following morning from participants who had fasted for at least 8 hours the previous night. We utilized Myriad kits and biochemical tests to assess several indicators including fasting blood glucose (FBG), glucose tolerance test (OGTT)-2 hours blood glucose, TC, triglycerides (TG), LDL-C, and high-density lipoprotein cholesterol (HDL-C). Additionally, we measured TSH, thyroid peroxidase antibodies (TPOAb), and thyroglobulin antibodies (TgAb) using a Cobas 601 analyzer (Roche Diagnostics, Switzerland). Finally, free thyroxine (FT4) levels were measured when TSH was elevated, whereas FT4 and free triiodothyronine (FT3) levels were assessed after TSH had decreased.

### Clinical Diagnosis Criteria

Thyroid disorders were defined based on prior research ^17^, and the specific diagnostic criteria are shown in **Supplementary Table S1**. Hypertension was identified based on a systolic blood pressure (SBP) of ≥ 140 mmHg and/or diastolic blood pressure (DBP) of ≥ 90 mmHg or a prior diagnosis of the condition ^18^. Indicators of diabetes included fasting blood glucose (FBG) levels of ≥ 7.0 mmol/L, 2-hour Oral Glucose Tolerance Test (OGTT) levels of ≥ 11.1 mmol/L, typical diabetic symptoms and random blood glucose levels of 11.1 mmol/L, or previously reported diagnosis of diabetes mellitus ^19,20^. Hyperlipidemia was determined by elevated levels of total cholesterol (TC) of ≥ 6.2 mmol/L (240 mg/dL), triglycerides (TG) of ≥ 2.3 mmol/L (200 mg/dL), low-density lipoprotein cholesterol (LDL-C) of ≥ 4.1 mmol/L (160 mg/dL), high-density lipoprotein cholesterol (HDL-C) < 1.0 mmol/L (40 mg/dL) ^21^. Hyperuricemia was defined as serum uric acid levels of > 420 mmol/L (7.0 mg/dL) in men and > 360 mmol/L (6.0 mg/dL) in women ^22,23^. Smokers were classified as participants who had smoked at least one cigarette per day over the previous 12 months.

### Statistical analysis

Continuous variables were presented as mean ± standard deviation when they satisfied a normal distribution, and non-normally distributed information was displayed as the median and interquartile range. Comparisons between groups were made by Student t-test or Wilcoxon rank sum test. Categorical variables were shown as proportions and compared by the chi-square test. Logistic regression analysis was utilized to explore potential influences, and a least absolute shrinkage and selection operator (LASSO) regression was applied to control for multicollinearity between variables. All statistical analyses and image visualization were conducted on R software (version 4.1.0). Two-sided *P* values less than 0.05 were considered statistically significant differences.

## Results

### Characteristics of participants

A total of 2272 individuals were recruited in this study, consisting of 1194 (52.55%) urban inhabitants and 1168 (51.41%) females. The median age was 43 years (ranging from 18 to 82 years), and 64.17% of the participants held a high school diploma or above. Farmers made up the largest proportion of the population (34.33%), and the majority (33.76%) of them earned between 10,000 and 50,000 RMB per year. Detailed baseline information is provided in **Table 1**. A single ailment was reported by 31.07% of the participants, while 23.46% assessed themselves as disease-free. As the number of diseases increased, the count of respondents decreased, as shown in **Figure 1**.

**Figure 1.**
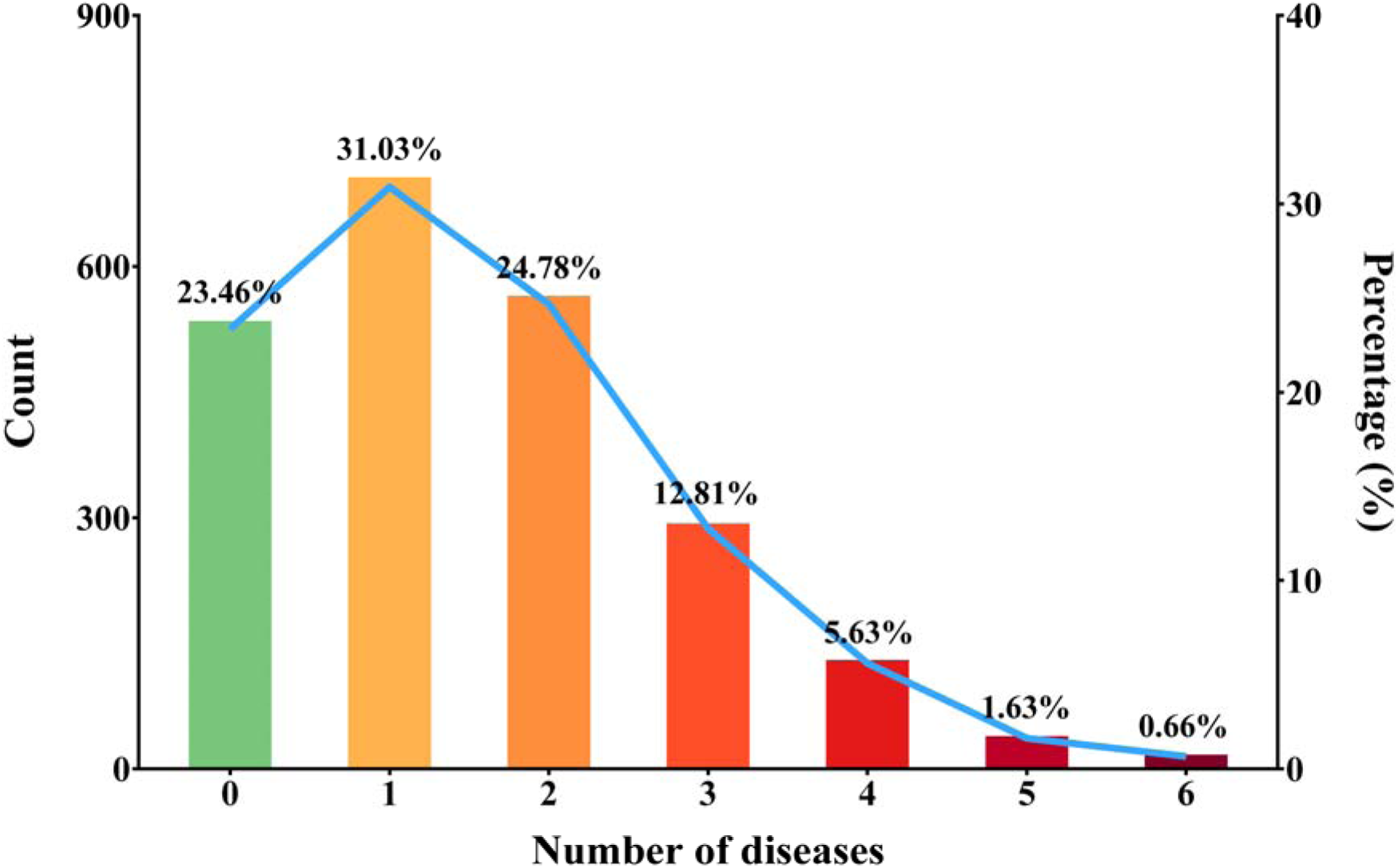
Count and percentage of participants suffering from different diseases.

**Table 1.**
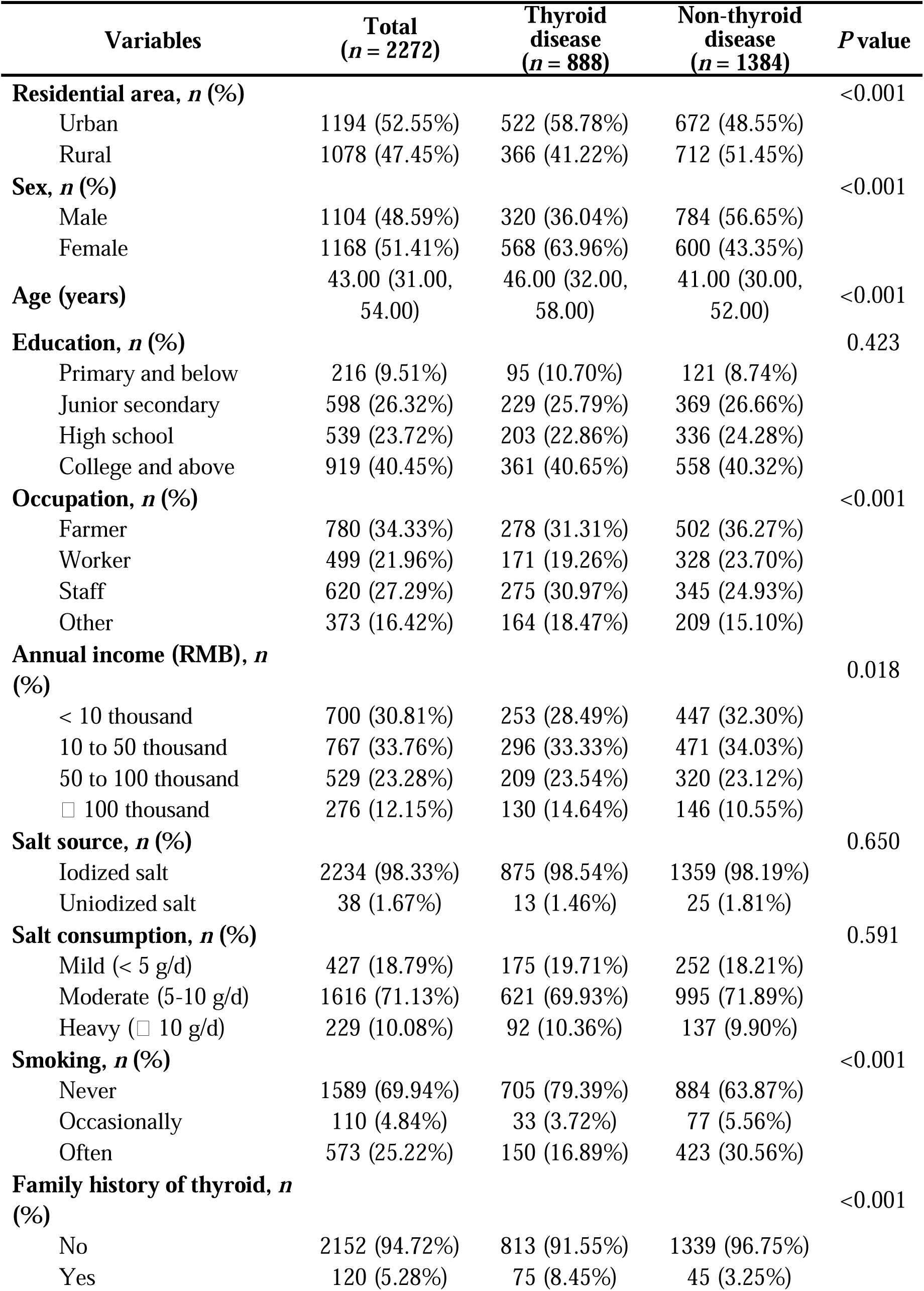
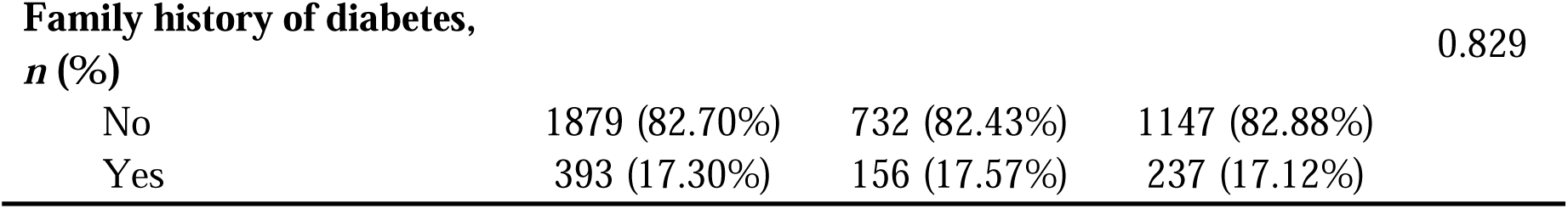
Demographic characteristics of participants.

### Prevalence of thyroid disorders

Of the 2272 study participants, 888 (39.08%) had thyroid diseases, including 33 (1.45%) with hyperthyroidism, 8 (0.35%) with subclinical hyperthyroidism, 49 (2.16%) with hypothyroidism, 408 (17.96%) with subclinical hypothyroidism, 395 (17.39%) with positive thyroid antibody, 285 (12.54%) with TPOAb positivity, 285 (12.46%) with TgAb positivity, 36 (1.58%) with goiter, and 246 (10.83%) with thyroid nodules. Notably, the prevalence of thyroid disorders was considerably higher among the urban-living participants (58.78% vs. 48.55%), with a staggering number of females (63.96% vs. 43.35%) affected. The study also indicated that those with a family history of thyroid disease suffered more (8.45% vs. 3.25%), and the aging population (46 vs. 41 years) was more susceptible. A detailed summary of these alarming findings can be found in **Table 1**.

### Comorbidity Combinations

The intricate relationship between thyroid diseases and concomitant conditions is a topic of great interest. In this study, among the 888 participants with thyroid diseases, a significant proportion of 62.50% had concurrent conditions. The most common comorbidities detected were hyperlipidemia, hypertension, diabetes, and hyperuricemia, with 41.22%, 29.05%, 16.44%, and 18.02%, respectively. The prevalence of having two concurrent conditions was 38.20%, and the number of participants with comorbidities decreased as the number of conditions increased. Notably, subclinical hypothyroidism and hyperlipidemia were the most frequent comorbidity pairing. The comprehensive listing of specific comorbidity combinations in **Supplementary Table S2** revealed an array of intriguing patterns. The top 10 most prevalent comorbidity combinations were further analyzed, revealing the top 3 dyad pairings to be subclinical hypothyroidism and hyperlipidemia (*n*=48), positive thyroid antibody and hyperlipidemia (*n*=26), and subclinical hypothyroidism and hypertension (*n*=25). Among triads of comorbidities, the subclinical hypothyroidism and positive thyroid antibody and hyperlipidemia (*n*=19) combination was most frequent, followed by subclinical hypothyroidism and hyperlipidemia and hyperuricemia (*n*=16), and subclinical hypothyroidism and hyperlipidemia and hypertension (*n*=15). Finally, the quadruple combination of thyroid nodule, diabetes, hypertension, and hyperlipidemia (*n*=11) emerged as the 9th most common (**Figure 2**).

**Figure 2.**
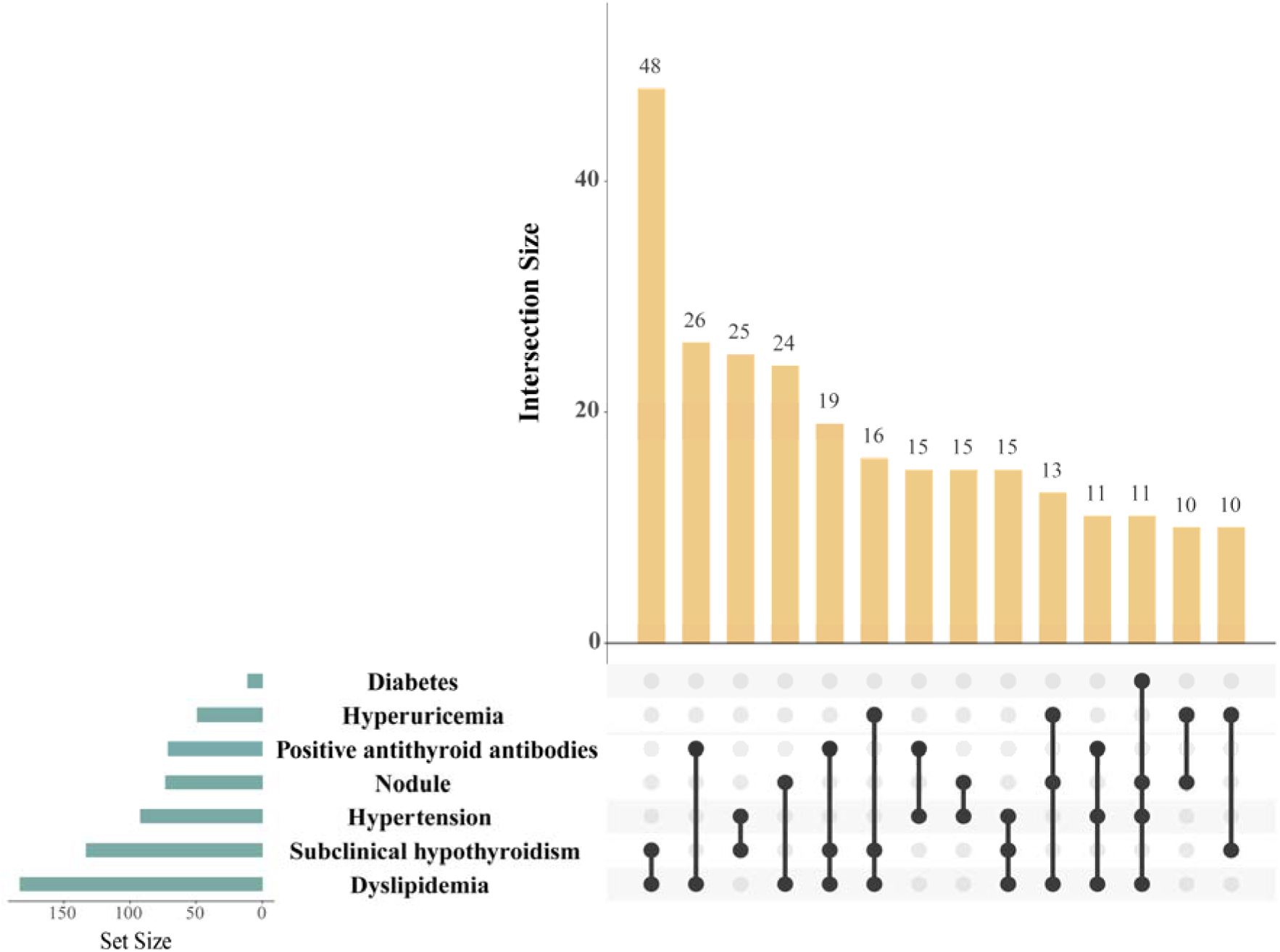
Top 10 combinations of the most prevalent comorbidity.

### Regression analysis of influencing factors for thyroid comorbidity

To investigate potential risk factors for comorbidity of thyroid disorder, the thyroid disease group was clustered according to whether they had any other comorbid conditions. The following variables were identified as being linked to thyroid disease comorbidity after a univariate logistic regression analysis: age, sex, education, occupation, salt consumption, iodine-containing food intake, smoking, family history of diabetes, weight, body mass index (BMI), waist circumference, and heart rate (**Table 2**). However, multicollinearity between the variables was detected and the findings indicated the presence of multicollinearity (**Supplementary Table S3**). In this case, LASSO regression was conducted to exclude the effect of covariates between variables, resulting in a more accurate assessment of the most significant risk factors (**Supplementary Fig. S3**). The results of LASSO regression suggest that female was a protective factor [odds ratio (OR)=0.55, 95% confidence interval (CI): 0.37-0.79, *P*=0.002], whereas older age (OR=1.06, 95% CI: 1.04-1.07, *P*<0.001), family history of diabetes (OR=1.76, 95% CI: 1.15-2.72, *P*=0.010), large waist circumference (OR=1.06, 95% CI: 1.04-1.08, *P*<0.001), and fast heart rate (OR=1.02, 95% CI: 1.01-1.04, *P*<0.001) were risk factors for thyroid disease comorbidity.

**Table 2.** Univariate and multivariate analysis of comorbidity.

| Variables | Univariate analysis |  | Multivariate analysis |  |
| --- | --- | --- | --- | --- |
|  | OR (95% CI) | *P* value | OR (95% CI) | *P* value |
| **Sex** |  |  |  |  |
| Male | 1 |  | 1 |  |
| Female | 0.38 (0.28, 0.51) | <0.001 | 0.55 (0.37, 0.79) | 0.002 |
| **Age** | 1.06 (1.05, 1.07) | <0.001 | 1.06 (1.04, 1.07) | <0.001 |
| **Education** |  |  |  |  |
| Primary and below | 1 |  |  |  |
| Junior secondary | 0.24 (0.13, 0.47) | <0.001 |  |  |
| High school | 0.36 (0.18, 0.71) | 0.003 |  |  |
| College and above | 0.15 (0.08, 0.28) | <0.001 |  |  |
| **Occupation** |  |  |  |  |
| Farmer | 1 |  |  |  |
| Worker | 0.80 (0.54, 1.19) | 0.274 |  |  |
| Staff | 0.80 (0.56, 1.13) | 0.210 |  |  |
| Other | 0.55 (0.37, 0.82) | 0.004 |  |  |
| **Salt consumption** |  |  |  |  |
| Mild (< 5 g/d) | 1 |  |  |  |
| Moderate (5-10 | 0.73 (0.51, 1.03) | 0.075 |  |  |
| Heavy (≥ 10 g/d) | 1.49 (0.84, 2.62) | 0.170 | | |
| **Iodine-containing food intake** |  |  |  |  |
| Never | 1 |  |  |  |
| Occasionally | 0.64 (0.40, 1.00) | 0.052 |  |  |
| Often | 0.79 (0.42, 1.47) | 0.455 |  |  |
| **Smoking** |  |  |  |  |
| Never | 1 |  |  |  |
| Occasionally | 1.39 (0.66, 2.91) | 0.383 |  |  |
| Often | 2.46 (1.63, 3.73) | <0.001 |  |  |
| **Family history of diabetes** |  |  |  |  |
| No | 1 |  | 1 |  |
| Yes | 1.54 (1.06, 2.24) | 0.023 | 1.76 (1.15, 2.72) | 0.010 |
| **Weight** | 1.05 (1.04, 1.07) | <0.001 |  |  |
| **BMI** | 1.24 (1.18, 1.30) | <0.001 |  |  |
| **Waist circumference** | 1.09 (1.07, 1.10) | <0.001 | 1.06 (1.04, 1.08) | <0.001 |
| **Heart rate** | 1.01 (1.00, 1.02) | 0.078 | 1.02 (1.01, 1.04) | <0.001 |

OR: odds ratio; CI, confidence interval; BMI: body mass index.

### Waist circumference differences in thyroid respondents in different sex

To further explore the relationship between waist circumference and the risk of thyroid disease comorbidity by sex, the waist circumference was categorized into different tertiles. As depicted in **Figure 3a**, the risk of thyroid disease comorbidity in males increased significantly with growing waist circumference, with a significantly higher risk observed in both the second (87-96 cm, OR=2.74, 95% CI: 1.51-5.07, *P*<0.01) and third (greater than 96 cm, OR=6.09, 95% CI: 2.96-13.64, *P*<0.001) tertiles compared to the first (less than 87 cm) tertile. Notably, a similar trend was observed in females, with a significantly higher risk observed in the second (77-86 cm, OR=2.38, 95% CI: 1.59-3.59, *P*<0.001) and third (greater than 86 cm, OR=6.21, 95% CI: 3.97-9.86, *P*<0.001) tertiles compared to the first (less than 77 cm) tertile (**Figure 3b**). It should be noted that the tertiles represent different ranges of waist circumference. These findings suggest that increased waist circumference is a significant risk factor for thyroid disease comorbidity in both male and female populations, with greater waist circumference associated with increased risk. These results further with greater waist circumference associated with increased risk. These results further highlight the importance of monitoring waist circumference as part of thyroid disease risk assessment.

**Figure 3.**
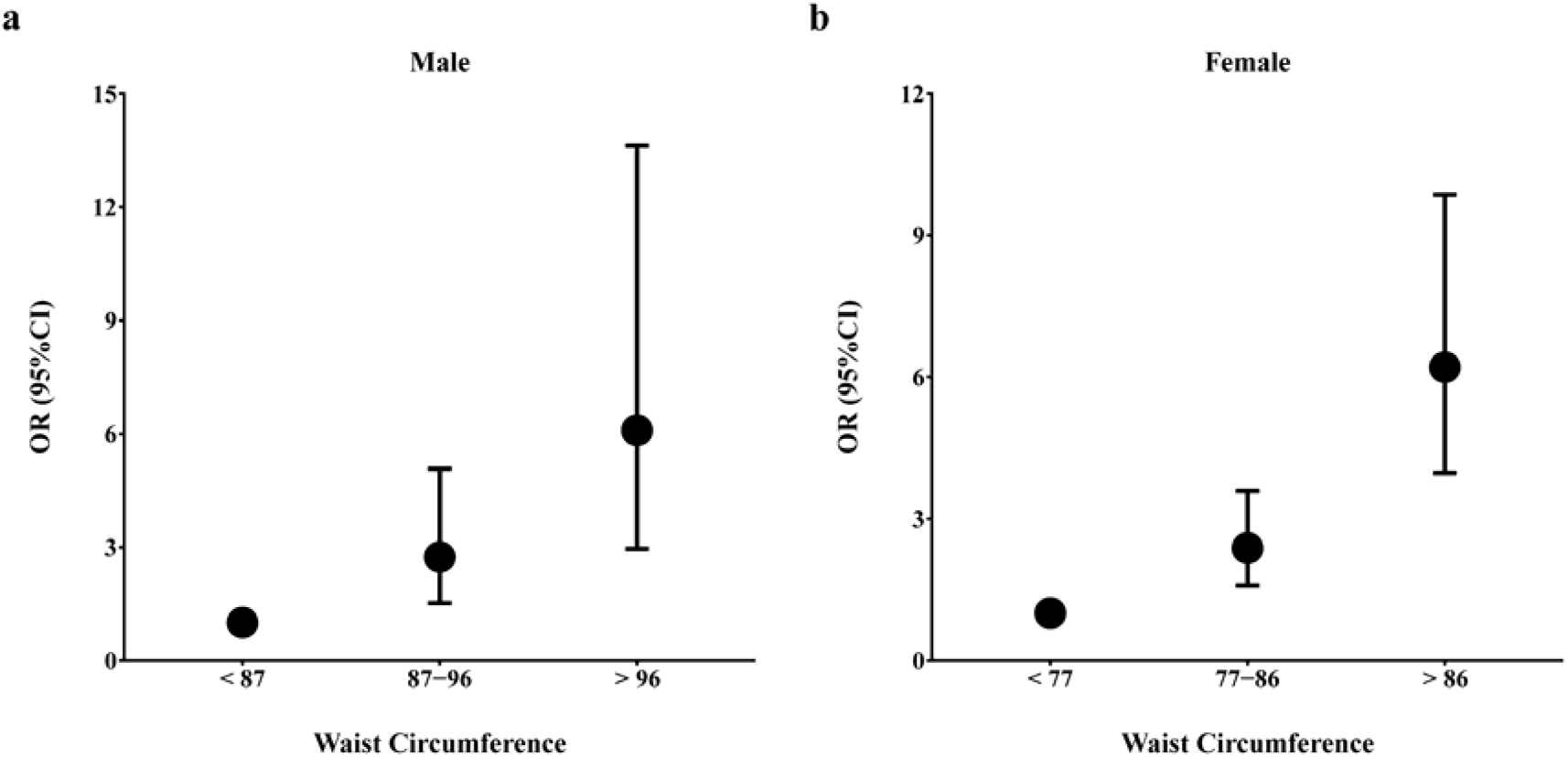
Waist circumference differences in thyroid respondents in different sex. (a) Male; (b) Female.

## Discussion

In this study, a total of 2272 participants were recruited to explore the patterns and associated factors of thyroid related disease comorbidity in Xi’an, China. The prevalence of thyroid disorders was staggering with 1.45% of hyperthyroidism, 0.35% of subclinical hyperthyroidism, 2.16% of hypothyroidism, 17.96% of subclinical hypothyroidism, 17.39% of positive thyroid antibody, 12.54% of TPOAb positivity, 12.46% of TgAb positivity, 1.58% of goiter, and 10.83% of thyroid nodule. Moving on to the prevalence rates of thyroid disorders, a nationally representative cross-sectional study from all 31 provincial regions of mainland China ^2^ revealed that the prevalence of hyperthyroidism, subclinical hyperthyroidism, hypothyroidism, subclinical hypothyroidism, positive thyroid antibodies, TPOAb positivity, TgAb positivity, goiter, and thyroid nodule in adults was 0.78%, 0.44%, 1.02%, 12.93%, 14.19%, 10.19%, 9.70%, 1.17%, and 20.43%, respectively. Except for subclinical hyperthyroidism and thyroid nodule, the prevalence rates of conditions in this study were greater than the national level. These rates suggest that thyroid problems are common in Xi’an, and more emphasis should be made on minimizing their frequency. While the underlying causes of higher thyroid disorder rates in Xi’an are not well-understood, several factors may be contributing to these trends. For example, population in Xi’an may have experienced an improving standard of living and overall quality of life in recent years, which could increase the prevalence of certain disorders. Additionally, differences in trace elements (such as iodine levels) ^17^, dietary habits ^24^, and economic levels across geographic regions may also play a role.

Hyperlipidemia is the most common comorbidity of thyroid disorders, with subclinical hypothyroidism and hyperlipidemia constituting the leading comorbidity pair. Previous research revealed a significant association between abnormal thyroid function and lipids ^25^. Additionally, individuals with clinical hypothyroidism had elevated levels of TC, TG, and LDL-C, whereas those with clinical hyperthyroidism had lower levels ^26,27^. It has also been discovered that TC, LDL-C, and TG increase with TSH even in people with normal thyroid function ^28^. Tognini et al. demonstrated that TSH was the second most important factor affecting TC, after age ^29^. The findings presented above support the occurrence of comorbid hyperlipidemia in thyroid disease. In addition, the proportion of different thyroid disease in this study varied, with the highest prevalence of subclinical hypothyroidism (17.96%), which may also explain the most frequent occurrence of hyperlipidemia in thyroid diseases. Therefore, thyroid disease control should be complemented by lipid screening and management to reduce comorbidity.

In addition, various risk factors for thyroid disease comorbidity were identified, including older age, family history of diabetes, large waist circumference, and fast heart rate, while the female was found to be a protective factor. In several investigations ^24,30^, age has been verified as a risk factor for thyroid conditions, which is consistent with the present study. While our study finds that women are a protective factor against thyroid disease comorbidity, previous investigations have suggested that women are more likely to develop thyroid problems ^24,30^, which may be due to the different study outcomes. The research mentioned above focused on the investigation of hypothyroidism and thyroid nodule, whereas our study spotlighted the exploration of thyroid related disease comorbidity. Although earlier research has indicated that BMI is linked to a higher risk of thyroid nodules ^31^, this investigation did not uncover a connection between BMI and thyroid disease comorbidity. BMI is a commonly used and valuable indicator of obesity but does not reveal lean body mass or central and peripheral body fat distribution ^32^. Waist circumference, a measure reflecting central obesity, was regarded to be more reliable than BMI in giving a precise picture of abdominal body fat. In addition, central obesity indices were considered to be more correlated with disease risk than BMI ^33–35^.

Through trichotomizing waist circumference in our study, we have conducted a rigorous statistical analysis and discovered a compelling correlation between waist circumference and the risk of thyroid disease comorbidity. Our findings are further supported by a study that investigated the relationship between thyroid function and metabolic syndrome, which revealed that an expanding waist circumference was associated with a higher risk of subclinical hypothyroidism ^36^. In addition, another independent research has also corroborated our results by demonstrating that an elevated waist circumference is positively linked to an increased prevalence of thyroid nodules ^37^. Taken together, these findings highlight the remarkable predictive power of waist circumference in assessing the risk of thyroid disease comorbidity. These results underscore the importance of recognizing the central role of waist circumference in evaluating the health status of individuals at risk of thyroid disease.

As with any scientific study, it is important to consider its limitations. In the case of this study, there are three primary areas of concern. Firstly, its cross-sectional design means that causality cannot be established with certainty. While the results suggest associations between certain risk factors and thyroid disease comorbidity, further research is needed to confirm these relationships. Secondly, the lack of information on respondents’ lifestyles is a potential confounding variable. Without a comprehensive understanding of factors such as diet, exercise, and drinking habits, it is difficult to fully explore the relationship between lifestyle and thyroid disease comorbidity. Finally, it is worth noting that the study only considered endocrine-related diseases as comorbidities. Further research is needed to explore the potential links between thyroid disease and other health indicators, such as cardiovascular disease or mental health.

Despite these limitations, however, this study sheds light on the high prevalence of thyroid-related disease comorbidity in the city of Xi’an. This highlights the need for more research into the underlying causes of this phenomenon and the potential preventive measures that could be taken. Ultimately, this study adds another piece to the ever-evolving puzzle of our understanding of thyroid disease and its comorbidities, and provides a foundation for future research in this important area.

## Data Availability

All data produced in the present study are available upon reasonable request to the authors

## Authors Contributions

YZ, TZ, BS designed the study. TZ and HL wrote the main manuscript text. YZ, XZ, HG, XL, ML, XD, and YS conducted the field survey. HL, ZG, WW, YY, and CY performed data analysis and prepared all the tables, figures, and supplementary materials for this manuscript. All authors reviewed the manuscript.

## Data availability statement

The datasets generated and/or analysed during the current study are not publicly available due to restrictions from the funding source and may be requested from the corresponding authors upon reasonable request.

## Acknowledgements

The authors thank all the participants and the doctors, nurses and technicians for their contributions and support in this study.

## Supplementary Materials

Supplementary Figure 1: Spatial distribution and survey area in China. The marked area represents Shaanxi Province and the star denotes the city of Xi’an. Supplementary Figure 2: Flowchart of participants recruitment. Supplementary Figure 3: Lasso regression model for variables selection. (a) Lasso coefficient profile with binomial deviation; (b) Distribution of lasso coefficients. Lasso: Least absolute shrinkage and selection operator. Supplementary Table 1: Diagnostic criteria for thyroid diseases. Supplementary Table 2: Count for the comorbidity conditions. Supplementary Table 3: Results of multicollinearity diagnostics. (*Supplementary Materials*)

