## Supplementary material for "Comorbidity Patterns and Associated Factors of thyroid related disorders: A community-based cross-sectional study in Chinese population": Supplementary Material.docx

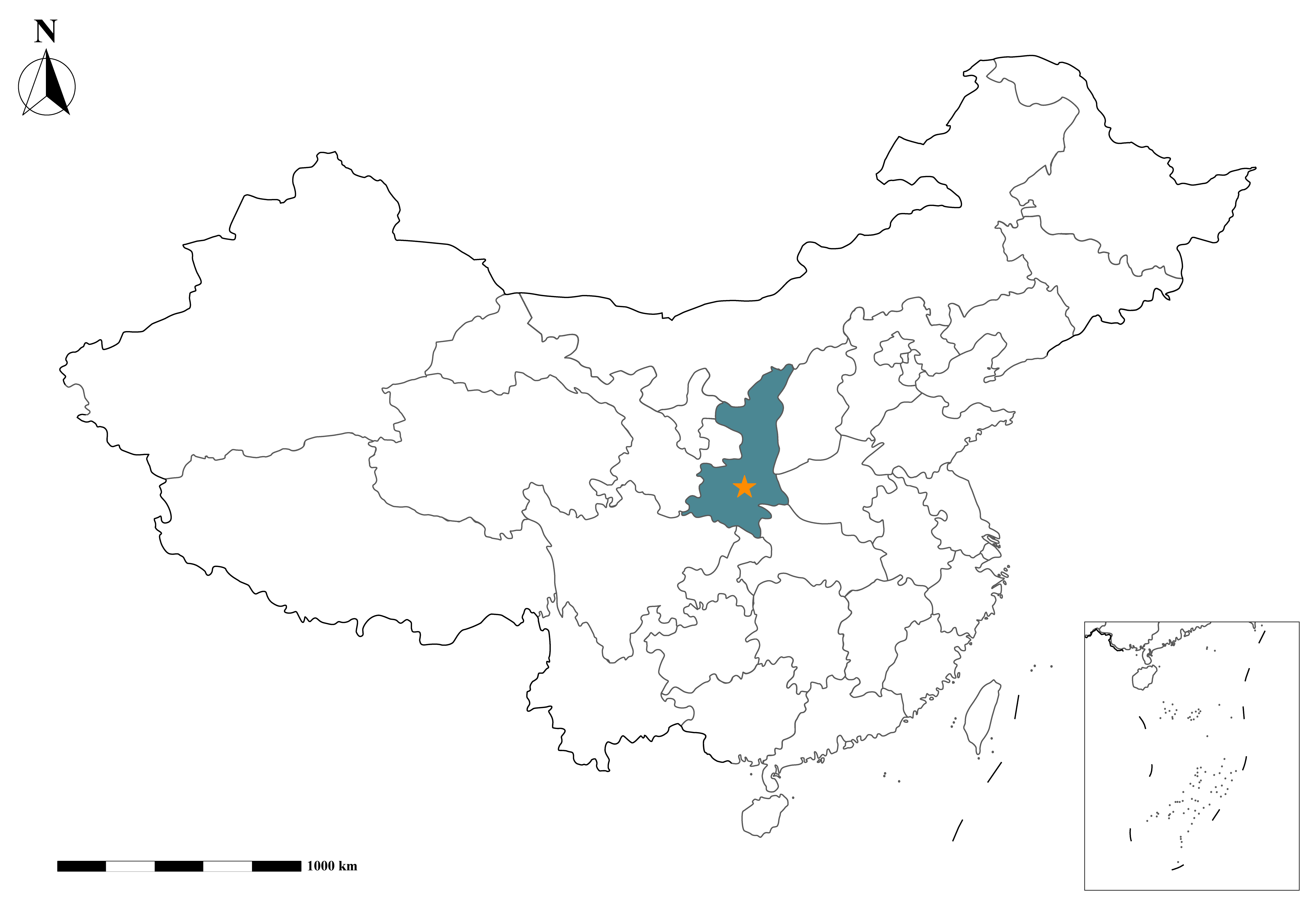

**Fig. S1** Spatial distribution and survey area in China. The marked area represents Shaanxi Province and the star denotes the city of Xi'an.

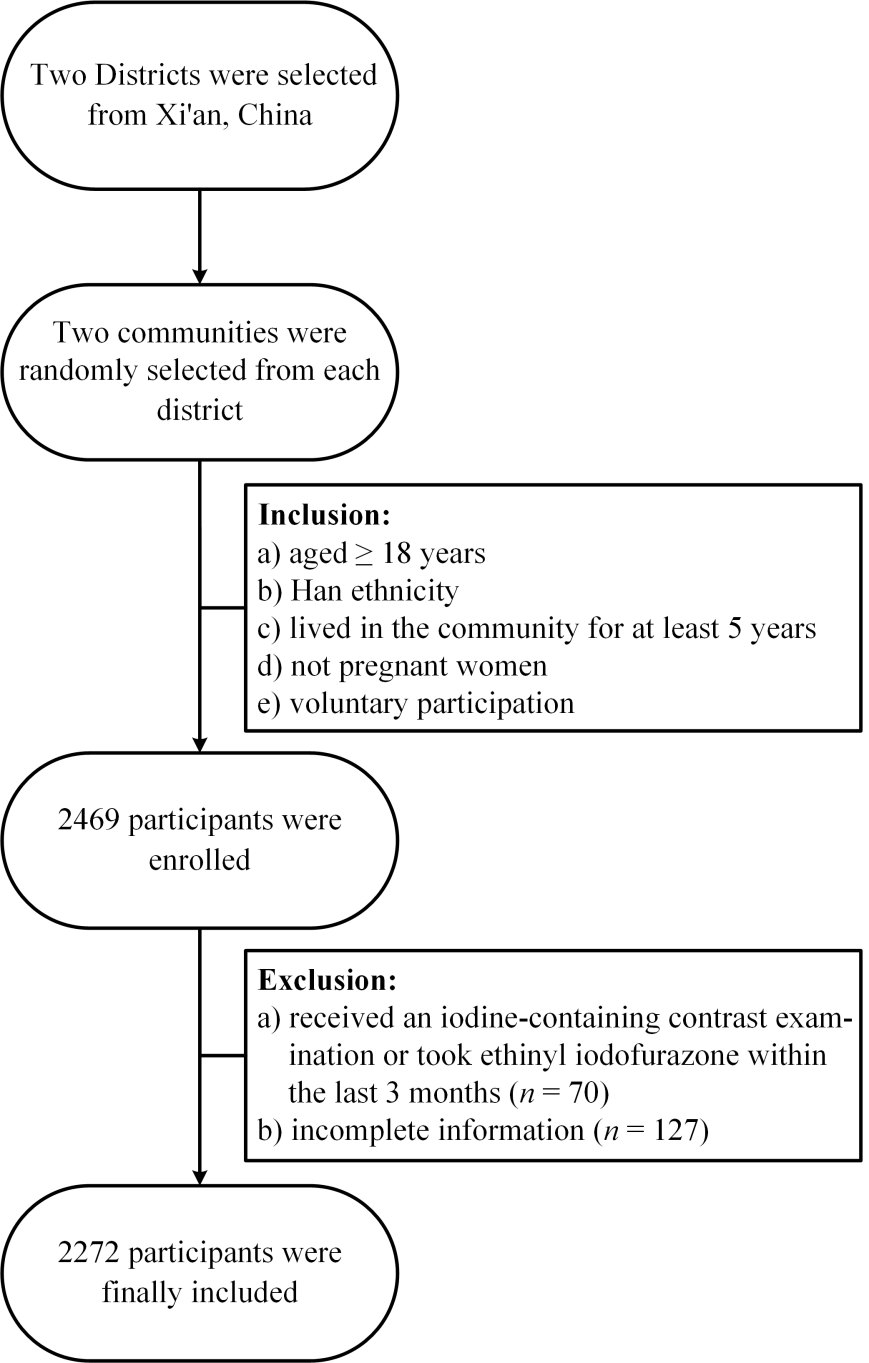

**Fig. S2** Flowchart of participants recruitment.

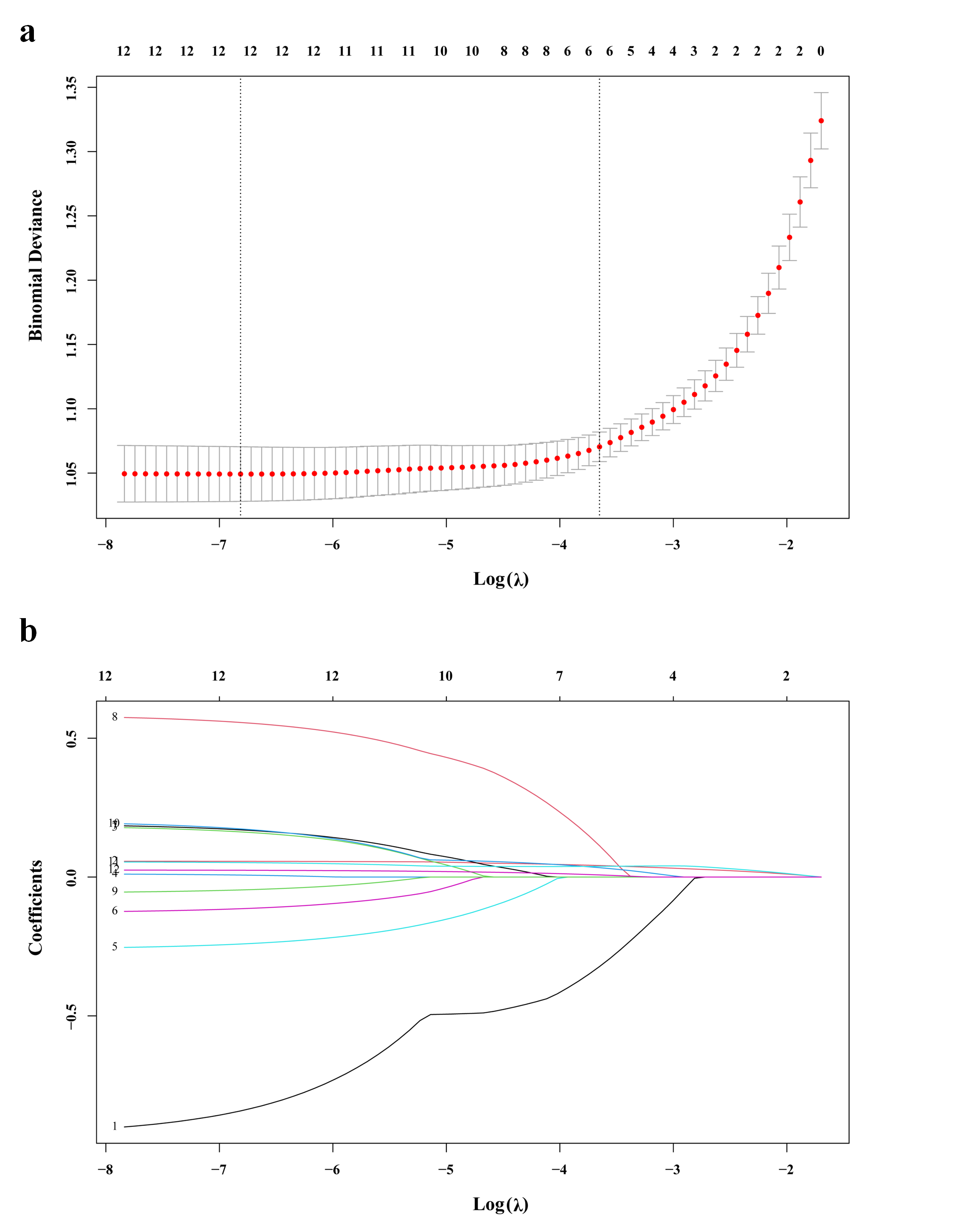

**Fig. S3** Lasso regression model for variables selection. **(a)** Lasso coefficient profile with binomial deviation; **(b)** Distribution of lasso coefficients. Lasso: Least absolute shrinkage and selection operator.

**Table S1** Diagnostic criteria for thyroid diseases.

| **Thyroid diseases** | **Diagnostic criteria** |
| --- | --- |
| Hyperthyroidism | TSH < 0.27 mIU/L, FT4 > 22.00 pmol/L, or FT3 > 6.80 pmol/L |
| Subclinical hyperthyroidism | TSH < 0.27 mIU/L, FT4: 12.00-22.00 pmol/L, and FT3: 3.10-6.80 pmol/L |
| Hypothyroidism | TSH > 4.20 mIU/L, FT4 < 12.00 pmol/L |
| Subclinical hypothyroidism | TSH > 4.20 mIU/L, FT4: 12.00-22.00 pmol/L |
| Positive thyroid antibody | TPOAb > 34 IU/mL or TgAb > 115 IU/mL |
| TPOAb positivity | TPOAb > 34 IU/ml |
| TgAb positivity | TgAb > 115 IU/ml |
| Goiter | Thyroid volume > 22.5 mL (men) or > 25.4 mL (women) |
| Thyroid nodule | One or more nodule (> 5 mm) without goiter |

TSH, thyroid stimulating hormone; FT4, free thyroxine; FT3, free triiodothyronine; TPOAb, thyroid peroxidase antibodies; TgAb, thyroglobulin antibodies.

**Table S2** Count for the comorbidity conditions.

| **Number of**  **conditions** | **Rank** | **Comorbidity conditions** | **Count** |
| --- | --- | --- | --- |
| 2 | 1 | Subclinical hypothyroidism and Dyslipidemia | 48 |
|  | 2 | Positive thyroid antibody and Dyslipidemia | 26 |
|  | 3 | Subclinical hypothyroidism and Hypertension | 25 |
|  | 4 | Thyroid Thyroid nodule and Dyslipidemia | 24 |
|  | 5 | Positive thyroid antibody and Hypertension | 15 |
|  | 6 | Thyroid Thyroid nodule and Hypertension | 15 |
|  | 7 | Subclinical hypothyroidism and Hyperuricemia | 10 |
|  | 8 | Thyroid Thyroid nodule and Hyperuricemia | 10 |
|  | 9 | Subclinical hypothyroidism and Diabetes | 9 |
|  | 10 | Thyroid Thyroid nodule and Diabetes | 6 |
|  | 11 | Positive thyroid antibody and Diabetes | 5 |
|  | 12 | Positive thyroid antibody and Hyperuricemia | 4 |
|  | 13 | Hypothyroidism and Dyslipidemia | 3 |
|  | 14 | Hypothyroidism and Hypertension | 3 |
|  | 15 | Hypothyroidism and Hyperuricemia | 3 |
|  | 16 | Goiter and Dyslipidemia | 2 |
|  | 17 | Goiter and Hypertension | 2 |
|  | 18 | Hyperthyroidism and Hypertension | 1 |
|  | 19 | Goiter and Hyperuricemia | 1 |
| 3 | 1 | Subclinical hypothyroidism and Positive thyroid antibody and Dyslipidemia | 19 |
|  | 2 | Subclinical hypothyroidism and Dyslipidemia and Hyperuricemia | 16 |
|  | 3 | Subclinical hypothyroidism and Dyslipidemia and Hypertension | 15 |
|  | 4 | Thyroid nodule and Dyslipidemia and Hyperuricemia | 13 |
|  | 5 | Positive thyroid antibody and Dyslipidemia and Hypertension | 11 |
|  | 6 | Subclinical hypothyroidism and Positive thyroid antibody and Hypertension | 9 |
|  | 7 | Subclinical hypothyroidism and Diabetes and Hypertension | 8 |
|  | 8 | Positive thyroid antibody and Diabetes and Dyslipidemia | 7 |
|  | 9 | Thyroid nodule and Dyslipidemia and Hypertension | 7 |
|  | 10 | Subclinical hypothyroidism and Thyroid nodule and Dyslipidemia | 6 |
|  | 11 | Subclinical hypothyroidism and Diabetes and Dyslipidemia | 6 |
|  | 12 | Subclinical hypothyroidism and Thyroid nodule and Hypertension | 5 |
|  | 13 | Positive thyroid antibody and Thyroid nodule and Dyslipidemia | 5 |
|  | 14 | Thyroid nodule and Hypertension and Hyperuricemia | 5 |
|  | 15 | Hypothyroidism and Positive thyroid antibody and Dyslipidemia | 4 |
|  | 16 | Subclinical hypothyroidism and Positive thyroid antibody and Hyperuricemia | 4 |
|  | 17 | Subclinical hypothyroidism and Hypertension and Hyperuricemia | 4 |
|  | 18 | Positive thyroid antibody and Diabetes and Hypertension | 4 |
|  | 19 | Thyroid nodule and Diabetes and Dyslipidemia | 4 |
|  | 20 | Hyperthyroidism and Positive thyroid antibody and Dyslipidemia | 3 |
|  | 21 | Hyperthyroidism and Positive thyroid antibody and Hyperuricemia | 3 |
|  | 22 | Subclinical hypothyroidism and Positive thyroid antibody and Diabetes | 3 |
|  | 23 | Positive thyroid antibody and Dyslipidemia and Hyperuricemia | 3 |
|  | 24 | Hypothyroidism and Positive thyroid antibody and Hypertension | 2 |
|  | 25 | Hypothyroidism and Dyslipidemia and Hypertension | 2 |
|  | 26 | Positive thyroid antibody and Thyroid nodule and Hypertension | 2 |
|  | 27 | Positive thyroid antibody and Thyroid nodule and Hyperuricemia | 2 |
|  | 28 | Goiter and Diabetes and Hypertension | 2 |
|  | 29 | Hyperthyroidism and Goiter and Hypertension | 1 |
|  | 30 | Subclinical hyperthyroidism and Positive thyroid antibody and Dyslipidemia | 1 |
|  | 31 | Subclinical hyperthyroidism and Thyroid nodule and Dyslipidemia | 1 |
|  | 32 | Hypothyroidism and Positive thyroid antibody and Diabetes | 1 |
|  | 33 | Hypothyroidism and Positive thyroid antibody and Hyperuricemia | 1 |
|  | 34 | Hypothyroidism and Diabetes and Dyslipidemia | 1 |
|  | 35 | Positive thyroid antibody and Thyroid nodule and Diabetes | 1 |
|  | 36 | Positive thyroid antibody and Hypertension and Hyperuricemia | 1 |
|  | 37 | Goiter and Dyslipidemia and Hypertension | 1 |
|  | 38 | Thyroid nodule and Diabetes and Hypertension | 1 |
|  | 39 | Thyroid nodule and Diabetes and Hyperuricemia | 1 |
| 4 | 1 | Thyroid nodule and Diabetes and Dyslipidemia and Hypertension | 11 |
|  | 2 | Subclinical hypothyroidism and Dyslipidemia and Hypertension and Hyperuricemia | 9 |
|  | 3 | Subclinical hypothyroidism and Diabetes and Dyslipidemia and Hypertension | 8 |
|  | 4 | Thyroid nodule and Dyslipidemia and Hypertension and Hyperuricemia | 8 |
|  | 5 | Subclinical hypothyroidism and Positive thyroid antibody and Dyslipidemia and Hypertension | 6 |
|  | 6 | Positive thyroid antibody and Dyslipidemia and Hypertension and Hyperuricemia | 4 |
|  | 7 | Hyperthyroidism and Positive thyroid antibody and Dyslipidemia and Hypertension | 3 |
|  | 8 | Subclinical hypothyroidism and Positive thyroid antibody and Goiter and Dyslipidemia | 3 |
|  | 9 | Subclinical hypothyroidism and Positive thyroid antibody and Thyroid nodule and Dyslipidemia | 3 |
|  | 10 | Subclinical hypothyroidism and Positive thyroid antibody and Dyslipidemia and Hyperuricemia | 3 |
|  | 11 | Positive thyroid antibody and Thyroid nodule and Dyslipidemia and Hypertension | 3 |
|  | 12 | Positive thyroid antibody and Diabetes and Dyslipidemia and Hypertension | 3 |
|  | 13 | Hypothyroidism and Positive thyroid antibody and Thyroid nodule and Dyslipidemia | 2 |
|  | 14 | Hypothyroidism and Positive thyroid antibody and Dyslipidemia and Hypertension | 2 |
|  | 15 | Subclinical hypothyroidism and Positive thyroid antibody and Diabetes and Dyslipidemia | 2 |
|  | 16 | Subclinical hypothyroidism and Thyroid nodule and Diabetes and Dyslipidemia | 2 |
|  | 17 | Subclinical hypothyroidism and Thyroid nodule and Dyslipidemia and Hypertension | 2 |
|  | 18 | Positive thyroid antibody and Diabetes and Dyslipidemia and Hyperuricemia | 2 |
|  | 19 | Thyroid nodule and Diabetes and Hypertension and Hyperuricemia | 2 |
|  | 20 | Hyperthyroidism and Positive thyroid antibody and Thyroid nodule and Dyslipidemia | 1 |
|  | 21 | Hyperthyroidism and Positive thyroid antibody and Thyroid nodule and Hypertension | 1 |
|  | 22 | Hyperthyroidism and Positive thyroid antibody and Diabetes and Dyslipidemia | 1 |
|  | 23 | Hyperthyroidism and Positive thyroid antibody and Diabetes and Hypertension | 1 |
|  | 24 | Hyperthyroidism and Diabetes and Dyslipidemia and Hypertension | 1 |
|  | 25 | Hyperthyroidism and Dyslipidemia and Hypertension and Hyperuricemia | 1 |
|  | 26 | Hypothyroidism and Positive thyroid antibody and Thyroid nodule and Diabetes | 1 |
|  | 27 | Hypothyroidism and Positive thyroid antibody and Diabetes and Dyslipidemia | 1 |
|  | 28 | Hypothyroidism and Positive thyroid antibody and Diabetes and Hyperuricemia | 1 |
|  | 29 | Hypothyroidism and Positive thyroid antibody and Dyslipidemia and Hyperuricemia | 1 |
|  | 30 | Hypothyroidism and Diabetes and Dyslipidemia and Hypertension | 1 |
|  | 31 | Subclinical hypothyroidism and Positive thyroid antibody and Thyroid nodule and Diabetes | 1 |
|  | 32 | Subclinical hypothyroidism and Positive thyroid antibody and Thyroid nodule and Hypertension | 1 |
|  | 33 | Subclinical hypothyroidism and Positive thyroid antibody and Thyroid nodule and Hyperuricemia | 1 |
|  | 34 | Subclinical hypothyroidism and Positive thyroid antibody and Diabetes and Hypertension | 1 |
|  | 35 | Subclinical hypothyroidism and Goiter and Diabetes and Dyslipidemia | 1 |
|  | 36 | Subclinical hypothyroidism and Thyroid nodule and Diabetes and Hypertension | 1 |
|  | 37 | Subclinical hypothyroidism and Thyroid nodule and Diabetes and Hyperuricemia | 1 |
|  | 38 | Subclinical hypothyroidism and Thyroid nodule and Dyslipidemia and Hyperuricemia | 1 |
|  | 39 | Subclinical hypothyroidism and Thyroid nodule and Hypertension and Hyperuricemia | 1 |
|  | 40 | Subclinical hypothyroidism and Diabetes and Dyslipidemia and Hyperuricemia | 1 |
|  | 41 | Subclinical hypothyroidism and Diabetes and Hypertension and Hyperuricemia | 1 |
|  | 42 | Positive thyroid antibody and Goiter and Diabetes and Dyslipidemia | 1 |
|  | 43 | Positive thyroid antibody and Thyroid nodule and Diabetes and Dyslipidemia | 1 |
|  | 44 | Positive thyroid antibody and Thyroid nodule and Dyslipidemia and Hyperuricemia | 1 |
|  | 45 | Positive thyroid antibody and Diabetes and Hypertension and Hyperuricemia | 1 |
|  | 46 | Goiter and Diabetes and Dyslipidemia and Hypertension | 1 |
|  | 47 | Goiter and Dyslipidemia and Hypertension and Hyperuricemia | 1 |
|  | 48 | Thyroid nodule and Diabetes and Dyslipidemia and Hyperuricemia | 1 |
| 5 | 1 | Subclinical hypothyroidism and Positive thyroid antibody and Diabetes and Dyslipidemia and Hypertension | 3 |
|  | 2 | Subclinical hypothyroidism and Positive thyroid antibody and Diabetes and Dyslipidemia and Hyperuricemia | 3 |
|  | 3 | Subclinical hypothyroidism and Diabetes and Dyslipidemia and Hypertension and Hyperuricemia | 3 |
|  | 4 | Goiter and Diabetes and Dyslipidemia and Hypertension and Hyperuricemia | 3 |
|  | 5 | Thyroid nodule and Diabetes and Dyslipidemia and Hypertension and Hyperuricemia | 3 |
|  | 6 | Subclinical hypothyroidism and Positive thyroid antibody and Goiter and Hypertension and Hyperuricemia | 2 |
|  | 7 | Subclinical hypothyroidism and Positive thyroid antibody and Dyslipidemia and Hypertension and Hyperuricemia | 2 |
|  | 8 | Positive thyroid antibody and Thyroid nodule and Diabetes and Dyslipidemia and Hypertension | 2 |
|  | 9 | Positive thyroid antibody and Thyroid nodule and Dyslipidemia and Hypertension and Hyperuricemia | 2 |
|  | 10 | Positive thyroid antibody and Diabetes and Dyslipidemia and Hypertension and Hyperuricemia | 2 |
|  | 11 | Hyperthyroidism and Positive thyroid antibody and Goiter and Dyslipidemia and Hyperuricemia | 1 |
|  | 12 | Hyperthyroidism and Positive thyroid antibody and Thyroid nodule and Dyslipidemia and Hyperuricemia | 1 |
|  | 13 | Subclinical hyperthyroidism and Positive thyroid antibody and Diabetes and Dyslipidemia and Hypertension | 1 |
|  | 14 | Hypothyroidism and Positive thyroid antibody and Goiter and Dyslipidemia and Hypertension | 1 |
|  | 15 | Hypothyroidism and Positive thyroid antibody and Diabetes and Dyslipidemia and Hyperuricemia | 1 |
|  | 16 | Hypothyroidism and Diabetes and Dyslipidemia and Hypertension and Hyperuricemia | 1 |
|  | 17 | Subclinical hypothyroidism and Positive thyroid antibody and Goiter and Diabetes and Dyslipidemia | 1 |
|  | 18 | Subclinical hypothyroidism and Positive thyroid antibody and Thyroid nodule and Diabetes and Hypertension | 1 |
|  | 19 | Subclinical hypothyroidism and Positive thyroid antibody and Thyroid nodule and Dyslipidemia and Hypertension | 1 |
|  | 20 | Subclinical hypothyroidism and Positive thyroid antibody and Diabetes and Hypertension and Hyperuricemia | 1 |
|  | 21 | Subclinical hypothyroidism and Goiter and Diabetes and Dyslipidemia and Hypertension | 1 |
|  | 22 | Subclinical hypothyroidism and Thyroid nodule and Dyslipidemia and Hypertension and Hyperuricemia | 1 |
|  | 23 | Positive thyroid antibody and Thyroid nodule and Diabetes and Dyslipidemia and Hyperuricemia | 1 |
|  | 24 | Positive thyroid antibody and Thyroid nodule and Diabetes and Hypertension and Hyperuricemia | 1 |
| 6 | 1 | Subclinical hypothyroidism and Positive thyroid antibody and Diabetes and Dyslipidemia and Hypertension and Hyperuricemia | 3 |
|  | 2 | Subclinical hypothyroidism and Thyroid nodule and Diabetes and Dyslipidemia and Hypertension and Hyperuricemia | 2 |
|  | 3 | Positive thyroid antibody and Thyroid nodule and Diabetes and Dyslipidemia and Hypertension and Hyperuricemia | 2 |
|  | 4 | Hyperthyroidism and Positive thyroid antibody and Diabetes and Dyslipidemia and Hypertension and Hyperuricemia | 1 |
|  | 5 | Hypothyroidism and Positive thyroid antibody and Goiter and Diabetes and Dyslipidemia and Hypertension | 1 |
|  | 6 | Hypothyroidism and Positive thyroid antibody and Goiter and Dyslipidemia and Hypertension and Hyperuricemia | 1 |
|  | 7 | Hypothyroidism and Positive thyroid antibody and Thyroid nodule and Dyslipidemia and Hypertension and Hyperuricemia | 1 |
|  | 8 | Subclinical hypothyroidism and Positive thyroid antibody and Thyroid nodule and Diabetes and Dyslipidemia and Hypertension | 1 |

**Table S3** Results of multicollinearity diagnostics.

| **Variables** | **VIF** |
| --- | --- |
| Gender | 2.813 |
| Age | 1.712 |
| Education | 2.924 |
| Occupation | 2.264 |
| Salt consumption | 1.117 |
| Iodine-containing food intake | 1.085 |
| Smoking | 1.807 |
| Family history of diabetes | 1.060 |
| Weight | 9.214 |
| BMI | 6.282 |
| Waist circumference | 3.367 |
| Heart rate | 1.090 |

VIF: variance inflation factor; BMI: body mass index.
